# British news media representations of mpox during the 2022 and 2024 outbreaks: a mixed-methods analysis using corpus linguistics

**DOI:** 10.1101/2025.09.30.25336981

**Authors:** Beth Malory, Marthe Le Prevost, Emily Jay Nicholls, Davide Bilardi, Aaron Choudhry, Shema Tariq

**Author notes:** Corresponding author Dr Beth Malory, Foster Court, Malet Place, Gower Street, London, WC1E 6BT, UK.

## Abstract

**Background:** Since 2022, over 100,000 people across 100 countries have been diagnosed with mpox (formerly monkeypox, renamed by the World Health Organization (WHO) in November 2022). News media plays a central role in outbreaks, disseminating information and shaping public discourse. Corpus linguistic approaches to massive language datasets can reveal how such outbreaks are represented but remain under-used in public health. Using these methods, we investigated representations of mpox in British news media during the 2022 and 2024 outbreaks.

**Methods:** We analysed the 83-billion-word English Trends corpus in SketchEngine, quantifying use of “*monkeypox*” and “*mpox”* in 2022–2024, and applying Corpus-Assisted Discourse Studies to compare news media content from 2022 and 2024 (peak incidence periods), comprising 1.2 billion and 500 million words, respectively. Using corpus linguistic tools, we explored the contexts in which *mpox* and *monkeypox* occurred, assessing shifts in representation.

**Findings:** Monthly use of “monkeypox” peaked at 0.07 occurrences per million words (n=6591) in May 2022, dropping by 99.8% by November 2022. Grammatical and lexical analysis of 2022 reporting found frequent attribution blame for transmission, particularly to gay and bisexual men who have sex with men (GBMSM). In 2024, coverage adopted more neutral language, largely avoiding stigmatisation.

**Interpretation:** UK news media reporting on mpox shifted from stigmatising language in 2022, often targeting GBMSM, to more neutral and inclusive coverage in 2024. The WHO-endorsed nomenclature change may have contributed, illustrating the impact of such interventions. This study demonstrates the value of corpus methods in tracking linguistic representations of infectious disease outbreaks.

**Funding:** This work is part of the VERDI project (101045989) which is funded by the European Union. Views and opinions expressed are however those of the authors only and do not necessarily reflect those of the European Union. BM, MLP and ST are partly funded through a Wellcome Accelerator Award held by ST (316319/Z/24/Z).

**Research in context:** *Evidence before this study:* We reviewed existing literature on media representations of mpox, and other infectious diseases, focusing on stigma, framing, and public perception. We searched academic databases, including studies that examined media discourse and linguistic framing, and those using qualitative or corpus-based methods. While some explored stigma in mpox media coverage, few studies applied computational (computer-based) linguistic analysis to large-scale media datasets, or compared media narratives across different outbreak periods.

*Added value of this study:* This study is the first to use a ‘big data’ approach to explore representations of mpox. It uses corpus linguistic methods to analyse over two billion words of British news media content across two mpox outbreaks. It reveals a shift from stigmatising language in 2022—often targeting specific communities—to more neutral and inclusive reporting in 2024. The findings demonstrate how media language evolves in response to public health guidance and highlight the potential of corpus linguistic methods to uncover patterns in public discourse.

*Implications of all the available evidence:* Media language plays a powerful role in shaping public understanding and attitudes during health emergencies. This study shows that changes in terminology and framing can reduce stigma and improve public health communication. These insights support the need for proactive media guidance and the use of linguistic analysis to inform future policy, practice, and research in outbreak response and health communication.

## Background

Mpox (formerly known as monkeypox), first identified in 1958, is a zoonotic disease caused by the monkeypox virus. Endemic in Central, East and West Africa since 1970, it remains a public health concern. Mpox has two clades: the more severe Clade I, reported in Central and East Africa, and the milder Clade II reported in West Africa.^1^

Mpox was declared a Public Health Emergency of International Concern (PHEIC) in May 2022 and again in August 2024.^2^ In May 2022, a global outbreak of the newly identified Clade IIb was first detected in the UK and spread to over 110 countries, particularly in Europe and North America. Later, in November 2022, the Word Health Organization endorsed a change in nomenclature to “mpox” to counteract rapidly emerging stigma and racism.^3^ By mid-2024, over 90,000 cases and more than 150 deaths were reported globally^4^ primarily among gay and bisexual men (GBMSM). Since 2023 a new Clade Ib variant has emerged, causing outbreaks in the Democratic Republic of the Congo (DRC) and neighbouring non-endemic countries, amidst ongoing Clade 1a and II outbreaks. As of August 2025, there had been 52,815 confirmed cases of mpox across Africa, and 229 deaths, in the preceding twelve months.^5^

The first UK case in the 2022 mpox outbreak was reported on 6 May 2022, with a total of 3,732 UK cases recorded by the end of 2022, peaking in mid-July.^6^ Before this outbreak, there had only been seven reported mpox cases in the UK, of which four were imported and three were among close contacts.^6^ Although case numbers in the UK have since declined, transmission in the UK continues, with 572 Clade IIb cases reported between 2023 and 31 July 2025. The decline is likely due to vaccine rollout amongst high-risk groups, providing protection.^7^

Print and digital news media play a crucial role during infectious disease outbreaks by disseminating information, conveying public health messaging, and shaping both individual perceptions and broader public discourse.^8^ Notably, Mpox emerged while COVID-19 was still having a global impact, and garnered significant media coverage. A UK-based survey in July 2022, using community social media and the Grindr dating app, found that 27% of 1,932 respondents reported learning about the mpox outbreak through printed press, highlighting its continued relevance as an information source.^9^ Despite this, limited research has engaged critically with how traditional news media portrayed the outbreak. Existing studies on media coverage have primarily focused on other infections, such as HIV and COVID-19,^8,10–12^ with little focus on how mpox has been represented, especially as an evolving public health crisis. To address this gap, this study utilises methods from corpus linguistics to analyse English language news media coverage of mpox.

Corpus linguistics is the study of language using large collections of real-world texts, known as corpora (singular, ‘corpus’).^13^ It offers powerful tools for analysing language use, revealing patterns of word associations, frequency, and context-specific meanings.^14^ For example, in an applied health research context, collocation analysis has been used to demonstrate that the word “*abortion”* is more likely to occur in anti-abortion contexts than neutral or pro-choice contexts.^15^ Despite its applicability, corpus linguistics remains underused in public health research, particularly in the study of media discourse during infection outbreaks, where it could offer valuable insights into how public health messages are framed and received. In this paper, we use corpus linguistic approaches to investigate representations of mpox in British news media during the 2022 and 2024 mpox outbreaks.

## Methods

This mixed-methods study uses corpus linguistic approaches to investigate representations of mpox between January 2022 and December 2024 in British news media (digital news articles, that provide news, commentary, and analysis on current events). This work is part of the broader VERDIQual study, which uses qualitative methods to understand mpox in Italy, Nigeria, Thailand and the UK.^16^

### Dataset

The 83-billion-word English Trends corpus is a regularly updated collection of English-language text from news articles and other online content, designed to monitor language use and changes over time. As a continuously updated dataset, it can provide timely insights into evolving infectious disease outbreaks. We accessed the English Trends corpus via SketchEngine,^17^ an online corpus management and text analysis interface. We compiled a 2.3-billion-word corpus of British news media outputs published between January 2022 and December 2024 for frequency analysis. We created sub-corpora for the calendar years 2022 and 2024, to compare linguistic framing of mpox during the outbreaks in those two years, corresponding to peaks in mpox incidence since 2022.

### Analysis

We undertook initial frequency analysis of the 2.3-billion-word corpus, quantifying frequency of “*monkeypox*” and “*mpox*” per calendar month between January 2022 and December 2024. Instances where *“monkeypox”* and *“mpox”* were used only to connote the virus were removed after manual checking to ensure we only captured data on discussions about *“monkeypox”* and *“mpox”* in relation to the disease outbreaks, since our focus was public perceptions of mpox the disease, rather than the virus. We tabulated the number and proportion of occurrences of each word by calendar month.

We used a combination of quantitative and qualitative methods to analyse the 2022 and 2024 sub-corpora separately, drawing on established methods from Corpus-Assisted Discourse Studies.^14^ First, we conducted *quantitative collocation analysis*, whereby we identified words which co-occurred with *“monkeypox”* or *“mpox”* (our “node words”) more frequently than would be expected by chance.^18^ This approach identifies strengths of association and patterns of usage, providing insights into how words acquire context-specific meaning.^15^ Collocation was measured using logDice, which minimises skewing of results when applied to very large datasets.^18^ Other parameters set for the collocation analysis included span size (the number of words to the left and right of a node word in the source articles) and the statistical threshold for logDice scores (a higher score representing a stronger relationship between node words and co-occurring word(s) (termed “collocates”). We applied a span size of 5, typical of such linguistic analyses,^19^ and a threshold logDice score of ≥2, with a minimum frequency of 5 instances of a collocate being used, allowing us to strike a balance between feasibility of manual analysis, and returning sufficient results for meaningful analysis. Where collocation is discussed, references to collocational strength refer to logDice score.

#### Key Corpus Linguistic Terminology

##### Collocates

words which are statistically proven to co-occur with a word of interest (also known as the **node word**, e.g. “*monkeypox*” or “*mpox*”) more often than would be expected by chance

##### LogDice

a measurement of collocation strength, where a higher score indicates a stronger relationship between the **node word** and its **collocate**

##### Span

the number of words either side of the **node word** that are considered when calculating the collocation statistic

Following collocation analysis, we conducted a qualitative *concordance analysis* of the 2022 and 2024 sub-corpora, whereby lines of text including the node words and collocates (termed *“concordance lines”)* are examined to understand the context in which they are used. We analysed a random sample of up to 100 concordance lines for *“monkeypox”* and *“mpox”* and collocates (or all co-occurrences, if a collocate occurred with a node word fewer than 100 times). Concordance lines were analysed thematically, using inductive coding, and analytic memos added to further develop understanding and broader themes.

All quantitative and qualitative analyses were conducted in SketchEngine.^17^

### Ethical approval

This analysis uses a dataset of news media available in the public domain. We do not use any individual-level data. Ethical approval was therefore not required.

## Results

### Frequency analysis

Use of “*monkeypox”* in British news media declined rapidly following the WHO’s endorsement of “*mpox”* as the preferred nomenclature in November 2022, in an effort to address stigma (Figure 1). Use of “*monkeypox”* in British news media peaked at 0.07 occurrences/million words of text in May 2022 (n=6591instances of the word), dropping to <0.01 occurrences/ million words (n=15) in November 2022; a 99.8% decrease in frequency. From November 2022 onwards, occurrences of “*monkeypox”* remained below 0.07 occurrences/million words for every subsequent month in our analysis.

**Figure 1.**
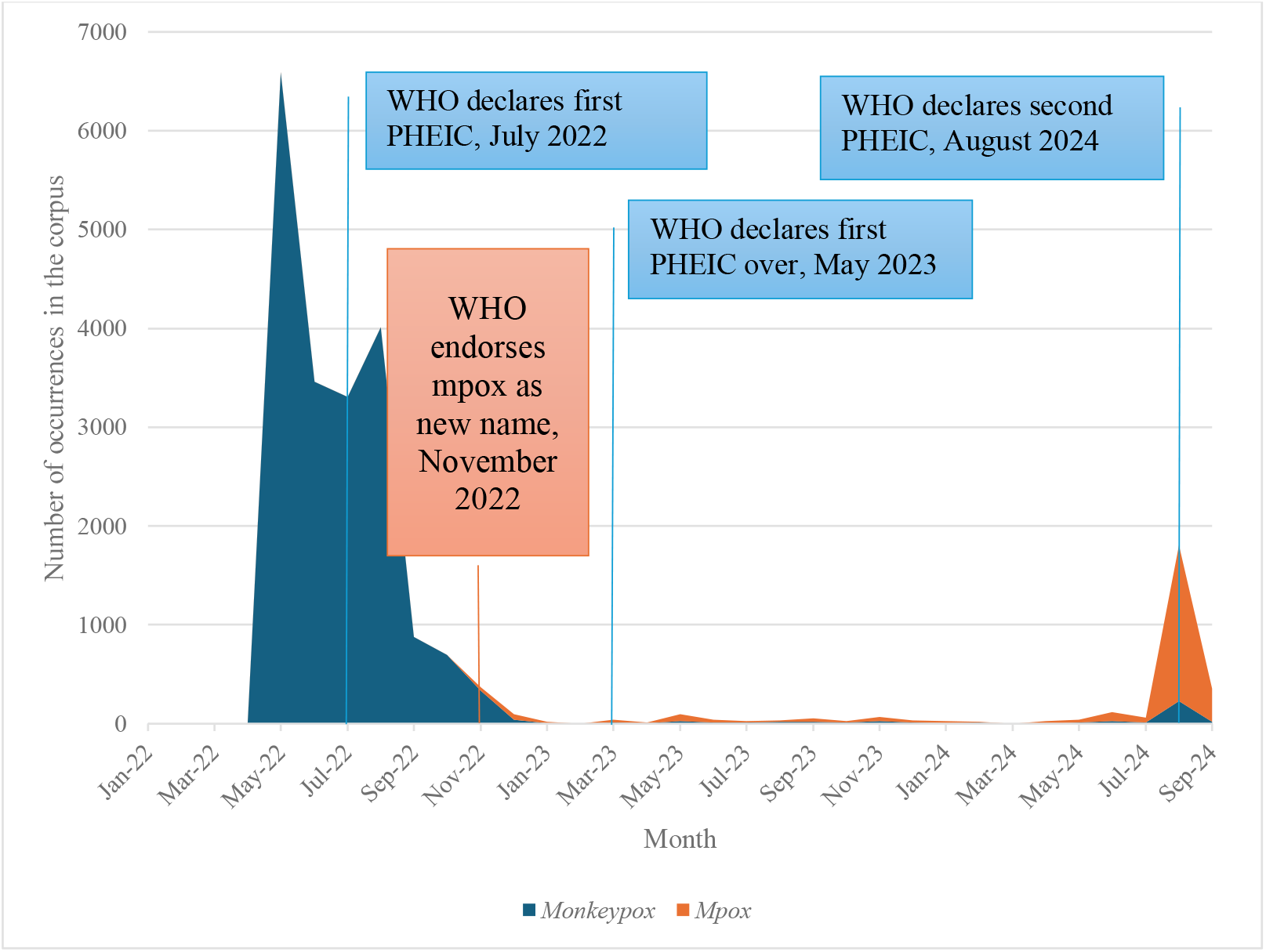
Occurrence of *monkeypox* and *mpox* in British news media (English Trends corpus).

The term “*mpox”* was first observed in British news media reports from November 2022 (Figure 1), corresponding to the WHO’s endorsement of it as the preferred nomenclature. However, “*mpox”* was used relatively infrequently between November 2022 and July 2024, demonstrating waning news media coverage as global incidence declined from late 2022. We observed a peak incidence of “*mpox”* in August 2024, reflecting the emergence of the Clade 1b outbreak and the designation of mpox as a PHEIC again in August 2024. However, this peak in use of “*mpox”* was substantially smaller than the initial peak in May 2022 at 0.02 occurrences/million words of text (1574 hits for *mpox* and 229 hits for *monkeypox)*. Combined, these 1803 occurrences account for only 23.4% of the total *monkeypox* occurrences in May 2022 (n=6591), showing clearly how much less newsworthy mpox was considered by August 2024.

We observed a clear shift in nomenclature usage, with “*mpox”* rapidly replacing “*monkeypox”* as the predominant linguistic variant in British news media, following the WHO’s endorsement in November 2022 (Figure 2). From November 2022 “*mpox”* occurred more frequently than “*monkeypox”* in nearly every month of our study period.

**Figure 2.**
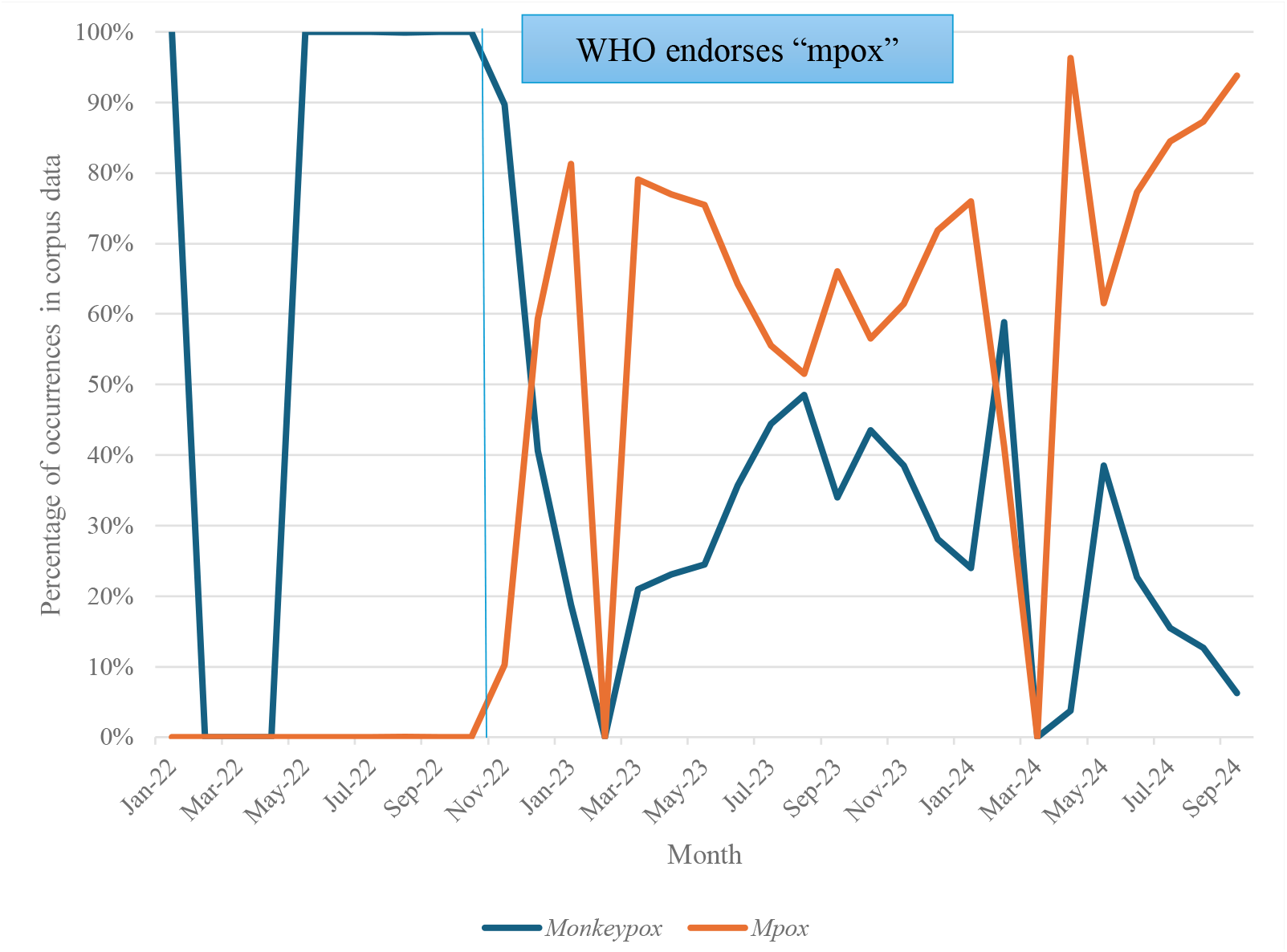
Percentage of mpox occurrences that were “*monkeypox”* versus “*mpox”* in the English Trends British news media sub-corpus, by calendar month.

### Collocation analysis

There were 161 collocates (words co-occurring more frequently with node words than expected) of “*monkeypox”* in the 2022 sub-corpus that met our threshold for inclusion (logDice score >2 and frequency >5). This indicated a strong association between each of these 161 words and “*monkeypox”*, which co-occured in a statistically significant way with each of these collocates, compared with its relationship with all other words in the sub-corpus. No collocates of “*mpox”* for 2022 met our inclusion thresholds, reflecting its adoption late in that year. We identified 47 and 6 collocates of “*mpox”* and “*monkeypox”* respectively in the 2024 sub-corpus. This smaller number of collocates reflects the substantially lower level of media coverage during this period.

### Concordance analysis

We further examined collocates identified in the 2022 and 2024 sub-corpora to determine how they were used alongside “*monkeypox”* or “*mpox”*, applying thematic analysis to concordance lines. In the 2022 sub-corpus, collocates found to co-occur strongly with *monkeypox* predominantly related to mode of transmission; 19.3% of the 161 collocates (n=31) appeared primarily or exclusively in conjunction with “*monkeypox”* in contexts describing transmission. Further analysis revealed that grammatical choices represented transmission of infection as either an active process, with an ‘agent’ responsible for transmission, or as a passive process without an identifiable ‘agent’. This is discussed in depth in later sections. The second largest set of codes for collocates of “*monkeypox”* in the 2022 sub-corpus related to symptoms and clinical presentation (12.4%, n=20). The third largest set related to classification i.e. viral clades (11.8%, n=19).

Our analysis of the 2024 sub-corpus revealed that the largest set of collocation codes associated with “*mpox”* were related to viral classification; 31.9% (n=15) of “*mpox”* collocates appeared primarily or exclusively in contexts discussing viral clades. The second-largest set of codes related to transmission (17.0%, n= 8), and the third largest to public health surveillance, i.e. monitoring and tracking the outbreak (12.8%, n=6). Analysis of the six “*monkeypox”* collocates identified in the 2024 sub-corpus revealed little about linguistic representation of mpox, often occurring when reporting the change in nomenclature (50.0%, n=3).

Whilst “*monkeypox”* and “*mpox”* in both the 2022 and 2024 sub-corpora often occurred in the context of discussion about transmission, patterns in how this was represented linguistically as either a passive or active process differed. In 2022, sentence construction subtly attributed responsibility to humans, rather than to the virus itself. The framing of “*monkeypox”* as the agent in a sentence i.e. the initiator of an action (in italics and underlined), was relatively rare in reporting of transmission in the 2022 sub-corpus:

*“<u>Monkeypox</u>* is *spreading* widely in North America and Europe in a way it never has before.”

(Pink News, 04/08/22)

In contrast, reference to human or animal agents were more prevalent in the 2022 sub-corpora:

*“<u>Anyone</u>* can get or *spread* monkeypox.*”*

(Pink News, 04/06/22)

Less commonly, sentences relating to transmission and including *monkeypox* in the 2022 sub-corpus, had no agent:

“It is not always clear how viruses such as *monkeypox* are *spread*.”

(*Manchester Evening News*, 04/05/22)

However, in other apparently agentless sentences, human or animal carriers of the virus (also underlined) are implied or explicitly stated:

“But *monkeypox* is mostly *spread* through sexual contact, at the moment particularly among *<u>men who have sex with men</u>.”*

(*Daily Mail*, 22/06/22)

“According to the NHS, *monkeypox* is *spread* by wild animals in parts of west or Central Africa”

(*Pink News*, 20/05/22)

These types of sentence construction encourage readers to see humans or animals (or groups thereof) as responsible for mpox transmission, without directly attributing transmission to these groups. A similar sentence structure was used with other verbs such as “*contract*” and “*transmit*”, further attributing responsibility for transmission to individuals, groups, or (less commonly) animals:

“Little is known about when *<u>he</u> contracted the monkeypox*.”

(*The Daily Mail*, 29/07/22)

“*<u>People</u>* can *transmit monkeypox* to their pets through close contact.”

(*Ex Bulletin*, 07/10/22)

The representation of mpox transmission as blameworthy in the 2022 sub corpus also extends to adjectives and nouns co-occurring with “*monkeypox”*. For instance, the adjective “*infected*” and the noun “*exposure”*, were both linked explicitly to sexual behaviour or identity:

“While *anyone* can become *infected* with *monkeypox*, the majority of cases continue to come from *<u>gay, bisexual and other men who have sex with men</u>* in interconnected sexual networks.”

(*Liverpool Echo*, 21/10/22)

*“*Some *<u>gay and bisexual men</u>*, who are at higher risk of *exposure* to *monkeypox* have been recommended to get the smallpox vaccine Imvanex.*”*

(*The Independent*, 30/07/22)

This association of mpox with sex, and particularly GBMSM, was a dominant theme in the British news media’s representation of mpox during the 2022 outbreak, reflecting the epidemiology of the outbreak in the UK at this time.

These two linguistic patterns (linking mpox transmission to active human or animal agents, and to GBMSM) were much less prominent in the 2024 sub-corpus. The largest set of collocate codes in 2024 related to viral classification (including “*clade*”, “*strain*”, “*1b*” and “*variant*”), rather than transmission. Reporting of mpox in 2024 British news media tended to be more neutral and factual than that observed in the 2022 sub corpus. Whilst 17.2% of the 47 collocates (n=8) for “*mpox”* in the 2024 sub-corpus related to transmission, the attribution of responsibility for transmission was less prevalent than in the 2022 sub-corpus. With *“spread”*, for example, 91.3% of occurrences of “*mpox”* lack a human or animal agent in 2024, by comparison with only 43.6% occurrences of “*monkeypox”* + “*spread”* in 2022.

Furthermore, in instances where mpox was discussed in the context of transmission, the focus was on information, often including reference to transmission in groups other than GBMSM:

“*Mpox* is primarily *spread* through close contact, such as sex, skin-to-skin contact and talking or breathing close to *<u>another person</u>*.”

(*The Telegraph*, 04/11/24)

“*<u>Pregnant women</u>* can also *spread mpox* to the foetus, during or after birth during skin-skin contact.”

(*The Telegraph*, 05/09/24)

Indeed, the above direct quotes were the only two instances of “*mpox”* co-occurring with “*spread” and* referring to or implying a human or animal agent. Instead, we observed that where “*spread”* co-occurred with “*mpox”* in the 2024 sub-corpus, “*mpox”* itself was the active agent in 91.3% (n=21) of instances, i.e. presenting mpox as actively transmitting *itself*, rather than ascribing responsibility for transmission to someone or something.

“*Mpox spreads* from person to person through close contact with someone who is infected”

(*BBC News*, 21/11/24)

Similarly, we note that verbs that co-occurred with “*monkeypox”* in the 2022 sub-corpus such as “*contract*” and “*infect*”, whilst also co-occurring with “*mpox”* in the 2024 sub-corpus, had been detached from blame and responsibility. Instead, “*contract”* was often used in relation to illness testimonies, often functioning to humanise the representation of infection. Likewise, “*infect”* occurred primarily in the context of public health advice; often using second-person pronouns such as “*you*” to address the reader directly, as opposed to using third-person pronouns to refer to some ‘other’:

“A *<u>man</u>* who *contracted mpox* has revealed how he *suffered* pus-filled acne and a hideous tonsil infection.”

(*The Mirror*, 06/09/24)

“If *<u>you</u>* get *infected* with *mpox*, it usually takes between five and 21 days for the first symptoms to appear, the NHS says.”

(*Express, 01/11/14*)

## Discussion

This study presents a novel application of corpus linguistic methods to a 2.3-billion-word dataset of British news media, revealing how language surrounding mpox has evolved since the start of the 2022 UK mpox outbreak. We describe how reporting shifted, at a linguistic level, from attribution of blame for its spread during the 2022 outbreak, to more neutral and destigmatised coverage by 2024.

Usage of *“monkeypox”* in British news media peaked in May 2022, declining to low levels by November of that year and remaining low from then on. “*Mpox*” emerged in British news media in November 2022, following the WHO’s endorsement of it as preferred nomenclature. Although its usage remained relatively low initially, a modest peak occurred in August 2024, coinciding with the Clade 1b outbreak and its classification as a PHEIC. Notably, this peak was substantially lower than the 2022 peak for *“monkeypox”*, indicating reduced media interest. This may reflect a combination of factors: fewer cases, geographic containment of the 2024 outbreak within the Africa region, and broader waning of interest in mpox.

The data also demonstrates widespread adoption of “*mpox”* by British news media, highlighting the influence of global public health bodies like the WHO in shaping media language in response to racialised and homophobic public discourse. This is in marked contrast to the pervasive use of stigmatising nomenclature in languages other than English.^20^ Our findings suggest that the change in English nomenclature may have contributed to fostering less stigmatising discourse.

In 2022, “*monkeypox*” collocates were predominantly linked to transmission, symptoms, and viral classification. By 2024, however, “*mpox*” was most frequently associated with viral classification, particularly relating to the emergence of Clade 1b, followed by transmission and public health surveillance. Mentions of *“monkeypox”* in the 2024 sub-corpus were limited, appearing mostly in discussions about nomenclature. This reflects the successful adoption of “*mpox*” in British news media and movement away from stigmatising narratives around mpox infection. Additionally, the prominence of viral classification, particularly references to variant clades, alongside a reduced emphasis on transmission, indicates a lower level of concern about domestic spread in the UK, given the geographical confinement of the Clade 1b outbreak within the Africa region. However, it is noteworthy that destigmatised coverage of mpox in 2024 British media reporting involves personal narratives and personal pronouns such as “you”, given the UK’s very low 2024 caseload. In this context of very low levels of infection in the UK, such direct address to the reader about potential infection, which was absent in the 2022 data, is surprising and likely reflects the destigmatisation of mpox infection between 2022 and 2024.

In both 2022 and 2024, transmission-related collocates were present, but the linguistic framing differed markedly. In 2022, sentence constructions often attributed responsibility to individuals or groups—through active grammatical agency and lexical associations with sexual activity, particularly among GBMSM. While reflecting epidemiological patterns, such language risks reinforcing stereotypes and perpetuating stigma. In contrast, 2024 reporting more frequently positioned mpox itself as the agent of transmission, rather than individuals. Verbs such as “*contract*” and “*infect*” were used mainly in the context of personal illness narratives or public health messaging, rather than implying responsibility.

To date, few studies have investigated news media reporting on mpox post-2022. Previous studies tended to focus on discursive representations and framing, such as comparisons to HIV or COVID-19, use of emotive language, and the role of advocacy in shaping narratives.^21^ Other work has explored how framing varies across countries and media platforms,^22^ and the media’s contribution to ongoing LGBTQ+ stigmatisation.^23,24^ Our study extends this literature by applying computational methods to massive language datasets, identifying how framing occurs at a linguistic level, and comparing two distinct outbreak periods. This allowed us to explore the dynamic nature of linguistic representation of an evolving infectious disease outbreak.

Language does not simply reflect reality, rather it helps construct it. It shapes public perception in subtle but powerful ways, through its presentation of agents, actions and affected groups. In this paper we demonstrate how, at a linguistic level, media discourse can reinforce stigma and assign blame during an infectious disease outbreak, whilst also potentially mitigating stigma through consistent use of neutral language, particularly when reinforced by guidance from major public institutions such as the WHO. These seemingly small shifts in grammar and vocabulary may have large effects on public attitudes, risk perception, and social responses. Our findings provide evidence that WHO guidance on mpox terminology did have the intended impact on public discourse in the UK. Corpus linguistic approaches, though currently under-used, hold substantial promise for understanding public discourse on infectious diseases and informing effective health communication strategies.

## Data Availability

All data produced are available at sketchengine.eu

